# Comparison of Simulated Outcomes Between Stool- and Blood-Based Colorectal Cancer Screening Tests

**DOI:** 10.1101/2022.10.27.22281611

**Authors:** A. Mark Fendrick, Vahab Vahdat, Jing Voon Chen, David Lieberman, Jordan J. Karlitz, Paul J. Limburg, A. Burak Ozbay, John B. Kisiel

## Abstract

**Objectives:** The Centers for Medicare & Medicaid Services (CMS) recommends covering blood-based tests meeting proposed minimum performance thresholds for colorectal cancer (CRC) screening. Outcomes were compared between currently available stool-based screening tests and a hypothetical blood-based test meeting CMS minimum thresholds.

**Methods:** Using the CRC-AIM model, outcomes were simulated for average-risk individuals screened between ages 45-75 years with triennial multi-target stool DNA (mt-sDNA), annual fecal immunochemical test (FIT), and annual fecal occult blood test (FOBT). Per CMS guidance, blood-based CRC screening was modeled triennially, with 74% CRC sensitivity and 90% specificity. Although not specified by CMS, adenoma sensitivity was set between 10-20%. Published adenoma and CRC sensitivity and specificity were used for stool-based tests. Adherence was set at (a) 100%, (b) 30-70%, in 10% increments, and (c) real-world rates for stool-based tests (mt-sDNA=65.6%; FIT=42.6%; FOBT=34.4%).

**Results:** Assuming perfect adherence, a blood-based test produced ≥19 lower LYG than stool-based strategies. At the best-case scenario for blood-based tests (100% adherence and 20% adenoma sensitivity), mt-sDNA at real-world adherence achieved more LYG (287.2 vs 297.1, respectively) with 14% fewer colonoscopies. At 100% blood-based test adherence and real-world mt-sDNA and FIT adherence, the blood-based test would require advanced adenoma sensitivity of 30% to reach the LYG of mt-sDNA (297.1) and approximately 15% sensitivity to reach the LYG of FIT (258.9).

**Conclusions:** This model suggests that blood-based tests with CMS minimally-acceptable CRC sensitivity and low advanced adenoma sensitivity will frequently yield inferior outcomes to stool-based testing across a wide range of adherence assumptions.

## Introduction

In 2021, there were an estimated 150,000 new cases of colorectal cancer (CRC) in the US and an estimated 53,000 CRC-related deaths.^1^ Fortunately, CRC screening has been demonstrated to effectively reduce disease-related incidence and mortality through earlier detection.^2, 3^ Broadly endorsed CRC screening options include relatively invasive tests, such as colonoscopy, that can identify and remove some colorectal neoplasia during the initial exam.^4-6^ Colonoscopies have high sensitivity and specificity to detect adenomas and CRC, but require bowel cleansing and often use sedation. Alternatively, non-invasive stool-based screening tests are taken at home without the pretest preparation or risks associated with colonoscopy (e.g. colon perforation). Stool-based tests, including fecal immunochemical test (FIT), fecal occult blood test (FOBT), and multi-target stool DNA (mt-sDNA), detect markers released from both CRCs and pre-cancerous adenomas^7^ and are recommended for average-risk screening by the US Preventive Services Task Force (USPSTF), American Cancer Society, and other major guidelines.^4-6^ All positive non-invasive stool-based screening tests should be followed up with colonoscopy.^4-6^ Blood-based biomarker tests represent a newer category of non-invasive CRC screening, but they are currently not recommended in major guidelines as first-line options.^4-6^

The Centers for Medicare & Medicaid Services (CMS) recommends covering blood-based biomarker tests with proposed minimum performance (sensitivity and specificity) thresholds for CRC screening test every 3 years for average-risk, asymptomatic Medicare beneficiaries ages 50–85 years.^8^ The proposed minimum performance thresholds are 74% for CRC sensitivity and 90% for CRC specificity compared with colonoscopy. These thresholds are based on the minimum sensitivity and specificity for FIT and mt-sDNA.^8^ Interestingly, the recently-issued CMS guidance does not stipulate performance thresholds for the detection of pre-cancerous adenomas, a critical outcome measure that substantially impacts the effectiveness of screening modalities.^8^

To provide further clarity regarding the potential clinical utility of blood-based CRC screening under a number of potential clinical scenarios, microsimulation modeling analyses were conducted to predict life-years gained (LYG), CRC incidence, and CRC mortality outcomes resulting from blood-based CRC screening based on CMS minimum thresholds. These outcomes were then compared with available stool-based screening options (FIT, FOBT, and mt-sDNA).

## Methods

### Microsimulation model

Colorectal Cancer and Adenoma Incidence and Mortality Microsimulation Model (CRC-AIM) is a previously validated microsimulation model that incorporates assumptions for the known natural history of CRC and for CRC screening test parameters to simulate the impact of screening on CRC outcomes (**Figure 1**).^9, 10^ The natural history component of the model follows the course of CRC using an adenoma-carcinoma sequence. The assumptions related to the natural history used in CRC-AIM have been previously described and include adenoma generation, adenoma growth, transition chance to pre-clinical CRC, CRC growth to become clinically detectable, and CRC death or survival, as described in **Figure 1**.^10^

**Figure 1.**
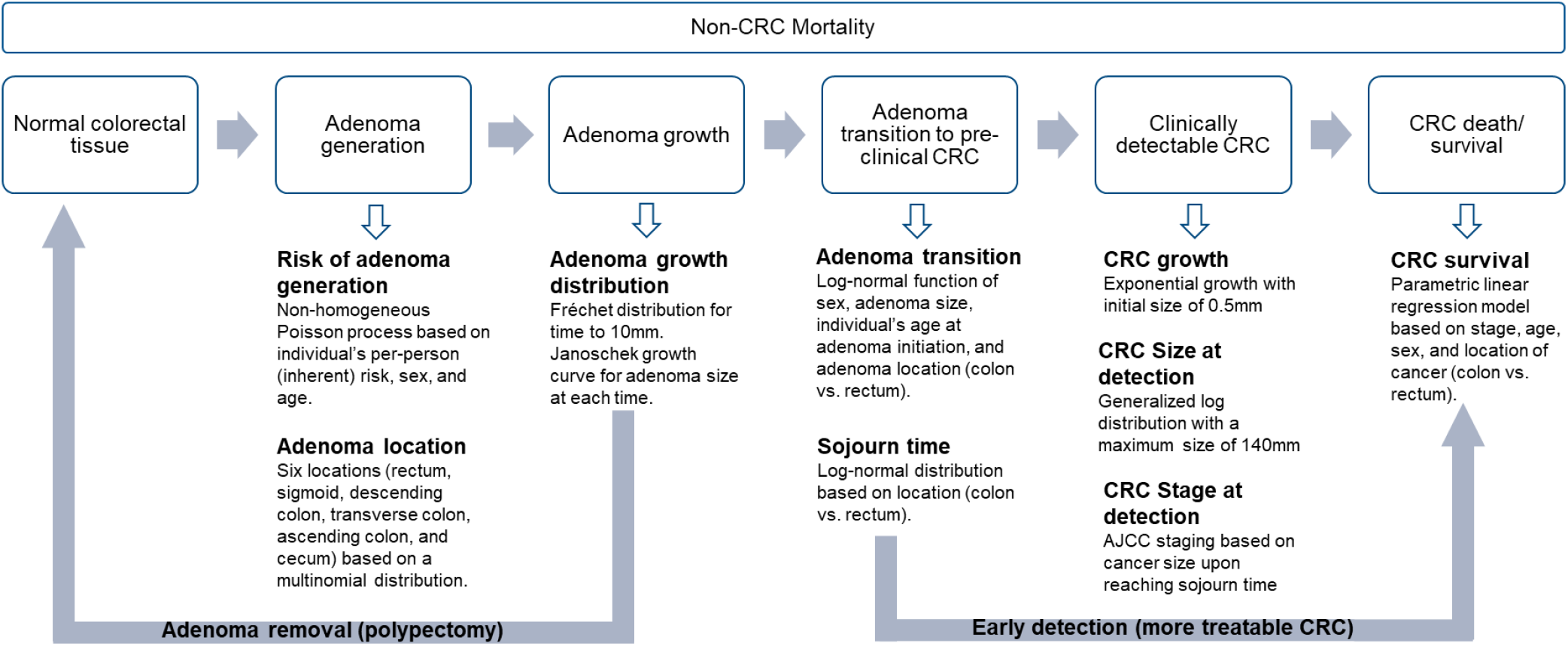
CRC-AIM microsimulation model. AJCC, American Joint Committee on Cancer; CRC, colorectal cancer.

The CRC screening test component of CRC-AIM incorporates sensitivity and specificity of each modality, the frequency each test is used, adherence to the assigned screening frequency, and age interval for screening.^9^ It is assumed that when a screening test is positive, either any identified pre-cancerous adenomas are removed to prevent the progression to preclinical CRC or that detection of early stage CRC is potentially more treatable, whereby mortality is reduced (**Figure 1**).

### Screening test performance assumptions

CRC-AIM was used to simulate outcomes of blood- and stool-based tests (FIT, FOBT, and mt-sDNA) for average-risk individuals free of diagnosed CRC at age 40 and screened between ages 45–75 years per USPSTF recommendations.^4^ Per CMS proposed criteria, CRC sensitivity and specificity for a blood-based test were set at 74% and 90%, respectively.^8^ Since CMS did not propose thresholds for adenoma sensitivity for a blood-based test, adenoma sensitivity was varied in 5 scenarios ranging from 10% to 20% (**Table 1**). The lower sensitivity of 10% is the false-positive rate of the CMS minimum threshold for specificity, and the maximum sensitivity of 20% is based on preliminary clinical results for a blood-based test.^11^ In two of the scenarios, adenoma sensitivity was assumed to be different for non-advanced (<10 mm) and advanced (≥10 mm) adenomas (**Table 1**). There is currently not enough evidence for blood-based test adenoma sensitivity based on adenoma size, so it was assumed that the adenoma detection for a non-advanced adenoma was half that of a large adenoma after excluding an accidental finding. Adenoma and CRC sensitivity and specificity for each stool-based test were those used by the USPSTF in their 2021 updated recommendations (**Table 1**).^12, 13^

**Table 1.** Screening test assumptions.

| Screening test, scenario | Sensitivity |  |  | Specificity | Screening Interval |
| --- | --- | --- | --- | --- | --- |
|  | Adenoma |  | CRC |  |  |
|  | <10 mm | ≥10 mm |  |  |  |
| mt-sDNA | 15%<sup>13</sup> | 42%<sup>13, a</sup> | 94%<sup>13</sup> | 91%<sup>13</sup> | 3 years |
| FIT | 7%<sup>13</sup> | 22%<sup>13, a</sup> | 74%<sup>13</sup> | 97%<sup>13</sup> | 1 year |
| FOBT | 5%<sup>12</sup> | 11%<sup>12</sup> | 68%<sup>12</sup> | 97%<sup>12</sup> | 1 year |
| Blood, S1 | 10% | 10% | 74%<sup>8</sup> | 90%<sup>8</sup> | 3 years |
| Blood, S2 | 12.5% | 15% | 74% | 90% | 3 years |
| Blood, S3 | 15% | 15% | 74% | 90% | 3 years |
| Blood, S4 | 15% | 20% | 74% | 90% | 3 years |
| Blood, S5 | 20% | 20% | 74% | 90% | 3 years |

CRC, colorectal cancer; FIT, fecal immunochemical test; FOBT, fecal occult blood test; mt-sDNA, multi-target stool DNA; S, scenario.
<sup>a</sup>Sensitivity for persons with advanced adenomas (i.e., ≥10 mm or adenomas with advanced history); sensitivity was not reported for the subset of persons with ≥10 mm adenomas separately from advanced adenomas.

### Screening test strategy assumptions

FIT and FOBT were recommended to be completed every year. The blood-based test and mt-sDNA were recommended to be completed every 3 years. For the primary analysis, adherence to screening intervals was assumed to be 100%. For secondary analysis, adherence was examined between 30–70%, in 10% increments, or at reported real-world adherence rates of 65.6% for mt-sDNA,^14^ 42.6% for FIT,^15^ and 34.4% for FOBT.^15^ Adherence to follow-up colonoscopies after a positive stool- or blood-based test was assumed to be 100%.

### Outcomes

Estimated outcomes were life-years gained (LYG) and reductions in CRC incidence and mortality compared with no screening. Outcomes were per 1000 individuals.

### Sensitivity analysis

To determine the threshold for adenoma sensitivity that would match outcomes with the stool-based tests, sensitivity analyses with greater than 20% non-advanced (<10 mm) and advanced (≥10 mm) adenoma sensitivities for the blood-based test were conducted.

## Results

In the scenario where a blood-based test meets CMS minimum performance thresholds on sensitivity and specificity and with perfect adherence assumed for all tests, a blood-based test produced ≥19 lower estimated LYG than currently available stool-based tests (**Figure 2**; **Supplemental Table S1**). When the *best-case* scenario for blood-based tests was modeled (100% adherence <u>and</u> 20% non-advanced and advanced adenoma sensitivity) and compared with real-world adherence rates for stool-based tests, mt-sDNA achieved more LYG (287.2 vs 297.1, respectively; **Figure 2**) with 14% fewer total colonoscopies (1601.9 vs 1404.5, respectively; **Supplemental Table S1**). High adherence (>60%) and high non-advanced and advanced adenoma sensitivity (both 20%) is needed for the blood-based test to exceed the LYG of FIT at real-world adherence (**Figure 2**) with 40% higher total colonoscopies (1409.7 vs 1004.9, respectively; **Supplemental Table S1**). Using real-world adherence for FOBT, the blood-test produces more LYG in most of the adherence and sensitivity scenarios (**Figure 2**).

**Figure 2.**
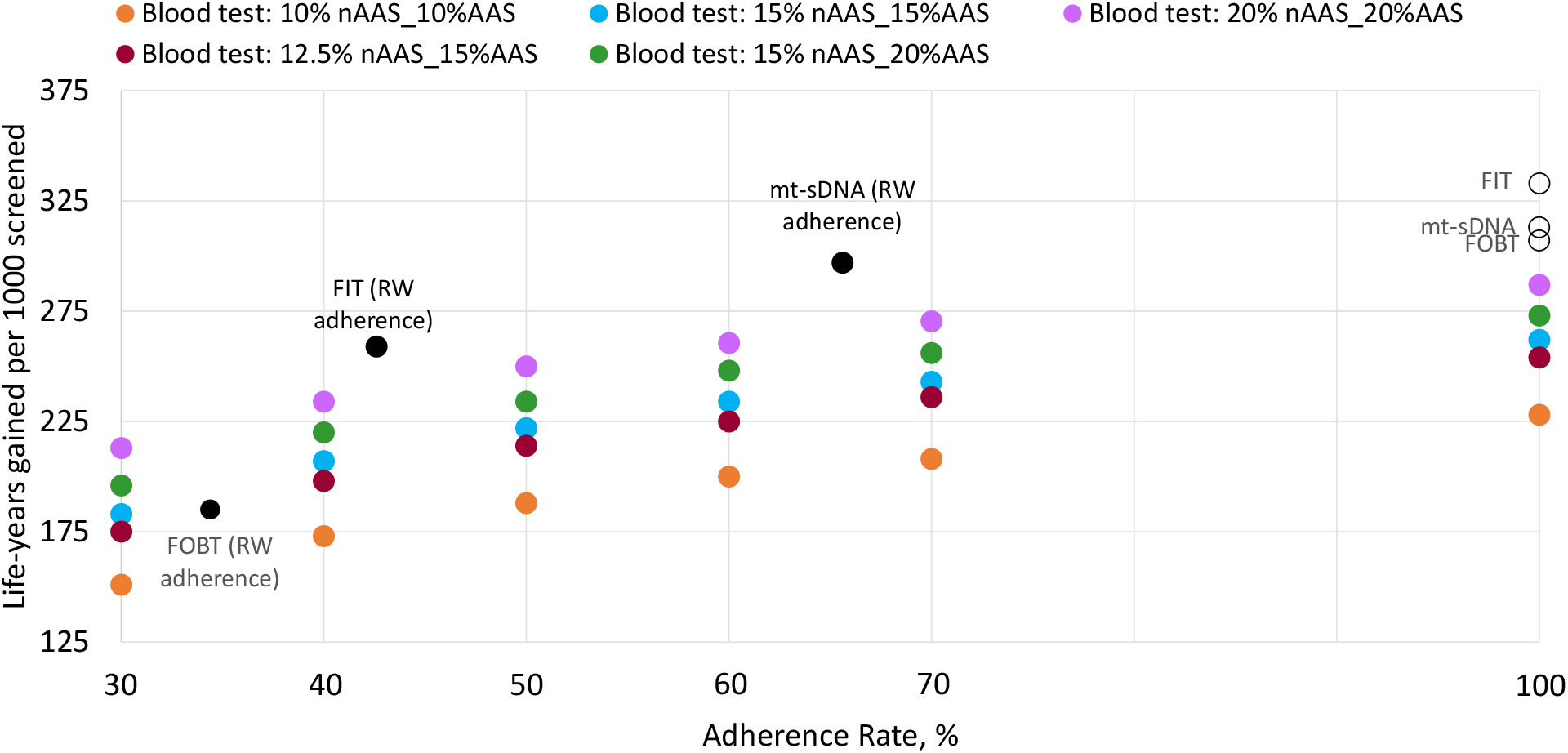
Life-years gained by adherence rate per 1,000 patients over lifetime horizon. AAS, advanced adenoma (≥10 mm) sensitivity; FIT, fecal immunochemical test; FOBT, fecal occult blood test; mt-sDNA, multi-target stool DNA; nAAS, non-advanced adenoma (<10 mm) sensitivity; RW, real-world.

Sensitivity analyses were conducted to determine what threshold of adenoma sensitivity would be required by a blood-based test to reach the same outcomes as mt-sDNA. At 100% adherence for the blood-based test and real-world adherence for mt-sDNA (65.6%) the blood-based test would require non-advanced adenoma sensitivity of 20% and advanced adenoma sensitivity of 30% to reach the LYG of mt-sDNA (297.1; **Supplemental Table S2)**. At real-world adherence for FIT (42.6%), approximately 15% sensitivity for both non-advanced and advanced adenomas would be required for the blood-based test to reach the LYG of FIT (258.9; **Supplemental Table S1)**.

**Figure 3** illustrates the tradeoffs in LYG produced between a blood-based test and each of the 3 stool-based screening tests across a broad range of adherence rates. In a scenario where a blood-based test has 10% sensitivity for both non-advanced (<10 mm) <u>and</u> advanced (≥10 mm) adenomas, mt-sDNA had at least 21 higher LYG (**Figure 3A**) and at least 11% and 7% higher reductions in absolute CRC incidence and mortality per 1000 individuals, respectively, at all explored adherence rates (**Supplemental Figures S1A and S2A**). When the blood-based test adenoma sensitivity for both small and advanced adenomas was increased to 20%, mt-sDNA and FIT had higher LYG (**Figure 3B**), and mt-sDNA had higher reductions in incidence and mortality, at identical adherence (**Supplemental Figures S1B and S2B**).

**Figure 3.**
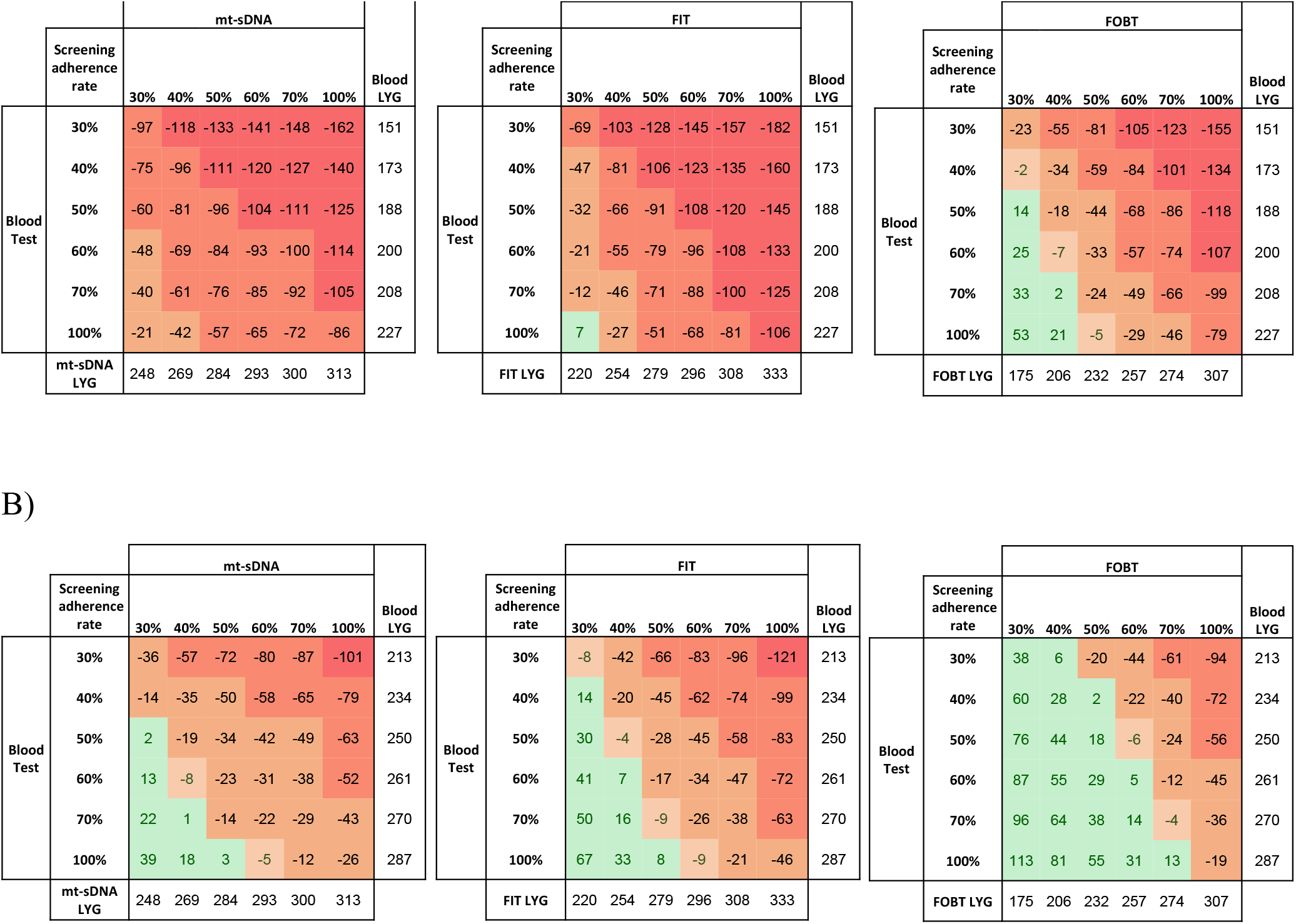
Life-years gained (LYG) per 1,000 patients screened over lifetime horizon based on different adherence rates for blood-based test adenoma sensitivity at A) 10% and B) 20%. Numbers represent ranges of incremental LYG versus the blood-based test. A positive value (green) indicates the stool-based test has less LYG than the blood-based test, and a negative value (red/orange) indicates the stool-based test has more LYG than the blood-based test. Adherence rates ranged from 100% or 30-70% for both stool- and blood-tests. FIT, fecal immunochemical test; FOBT, fecal occult blood test; mt-sDNA, multi-target stool DNA.

## Discussion

The results of this microsimulation model indicate that estimated LYG are inferior with a blood-based test that meets CMS minimum performance standards and interval testing recommendations as compared with currently endorsed, stool-based screening strategies when adherence to all tests is assumed to be 100%. Using real-world adherence rates for stool-based tests, mt-sDNA produced more LYG than the blood-based test with 100% adherence and best-case adenoma sensitivity. To exceed the LYG of FIT at real-world adherence, a blood-based test needed high adenoma detection and adherence. In contrast, the blood-based test produced more LYG than FOBT at real-world adherence.

The main driver of the scenarios where the blood-based test was inferior to the stool-based tests was the inability of the blood-based test to sufficiently detect advanced adenomas. Other models have also demonstrated the important contribution of the detection and removal of advanced adenomas in reducing CRC incidence and mortality.^16^ For this analysis, adenoma sensitivity for the blood-based test was assumed to be between 10-20% producing a wide range of LYG (10%, 228; 20%, 287) given perfect adherence. The sensitivity assumptions of the stool-based tests to detect adenomas (42% mt-sDNA, 22% FIT, and 11% FOBT) were the same as those used by the USPSTF in their 2021 recommendations^12^ and were derived from data compiled from clinical studies.^13^ In their assessment of blood-based tests for CRC screening,^8^ CMS identified 2 studies that reported advanced adenoma sensitivity.^17, 18^ These 2 studies used samples from the same patient population, but used first and second generation tests for which sensitivity was reported as 11% and 22%, respectively.^17, 18^ In addition, preclinical results released for another blood-based test reported an advanced adenoma sensitivity of 20%.^11^ Therefore, for the current analysis a lower limit for adenoma sensitivity was set at 10%, which is the false-positive rate of the CMS minimum threshold for specificity (essentially 0% adenoma sensitivity), and an additional range of adenoma sensitivities were applied up to a maximum of 20% to account for the full range of sensitivities at which the available blood-based tests are currently capable. The sensitivity analysis indicates that even when comparing the blood-based test at 100% adherence with mt-sDNA at real-world adherence, the blood-based test would require advanced adenoma sensitivity of 30% to achieve comparable clinical outcomes.

Screening for CRC needs to be repeated at intervals that vary depending on the screening test. Perfect adherence to the recommended 1- or 3-year intervals for non-invasive screening over the course of 30 years is not realistic in routine clinical practice. Thus, published real-world adherence rates of 65.6% for mt-sDNA,^14^ 42.6% FIT,^15^ and 34.4% for FOBT^15^ were applied in the model. Other adherence rates for the stool-tests have been published that vary by the population evaluated and how adherence was defined.^19-26^ Therefore, a range of adherence rates from 30% to 100% were also applied in the model to accommodate the potential variability in real-world adherence. Higher LYG with mt-sDNA was observed compared with the blood-based test (with 10% adenoma sensitivity) at all levels of adherence, even when assuming adherence as low as 30% for mt-sDNA and as high as 100% for the blood-based test. When the levels of adherence were identical between the tests, mt-sDNA and FIT maintained an advantage in LYG over the blood-based test at the highest assumed adenoma sensitivity (20% for non-advanced and advanced adenomas). The blood-based test at 20% adenoma sensitivity outperformed FOBT at lower adherence rates. The adherence to a triennial blood-based CRC screening test is unknown, but US self-reported screening surveillance data indicated that in 2009, 89% of adults aged 45-64 years in the general population had undergone blood cholesterol screening in the previous 5 years.^27^

A limitation of this analysis is that the sensitivity of the blood- and stool-based screening tests to detect sessile serrated polyps (SSPs) was not specifically modeled. SSPs develop from a different genetic pathway than the conventional adenoma pathway, and approximately 20-30% of CRCs arise from this alternate pathway.^28^ The sensitivity to detect SSPs varies among CRC screening tests. In one study, mt-sDNA was more sensitive than FIT to detect SSPs measuring ≥10 mm (42.4% vs 5.1%, respectively; P<0.001).^29^ The sensitivity of blood-based tests to detect SSPs is unknown. Another limitation is that the current analysis was limited to evaluation of a 3-year interval for blood-based tests, in alignment with the recently proposed CMS guidance. Assessment of additional intervals may be informative and could be considered for future analyses.

In sum, data from this modeling study suggest that blood-based screening strategies leveraging tests with minimally-acceptable sensitivity for CRC, coupled with low sensitivity for adenomas (using previously reported estimates), will yield clinical outcomes that are frequently inferior to stool-based strategies, even across a wide range of adherence assumptions. To realize meaningful public health benefits of CRC screening, strengths and weaknesses of various screening modalities must be conveyed to patients.

## Data Availability

All data produced in the present work are contained in the manuscript

## Acknowledgments

Medical writing and editorial assistance were provided by Erin P. Scott, PhD, of Maple Health Group, LLC, funded by Exact Sciences Corporation.

