## Supplemental Table for "Comparison of Simulated Outcomes Between Stool- and Blood-Based Colorectal Cancer Screening Tests"

### Supplemental Material

**Table S1**. Estimated outcomes with blood-based and stool-based test under various adherence and blood-based test adenoma sensitivity scenarios.

| Screening Test | Adherence | Total COLs | CRC cases | CRC deaths | LY with CRC | LYG | CRC incidence reduction | CRC mortality reduction |
|---|---|---|---|---|---|---|---|---|
| **No screening** | -- | 80.1 | 80.1 | 36.8 | 643.1 | 0.000 | 0.0% | 0.0% |
| **mt-sDNA** | **30%** | 1084.2 | 40.1 | 15.5 | 402.8 | 248.1 | 50.0% | 57.8% |
| | **40%** | 1208.4 | 36.6 | 13.9 | 375.1 | 269.1 | 54.3% | 62.2% |
| | **50%** | 1299.0 | 34.3 | 12.8 | 356.2 | 284.1 | 57.2% | 65.3% |
| | **60%** | 1371.7 | 32.6 | 12.0 | 343.1 | 292.6 | 59.3% | 67.4% |
| | **70%** | 1428.2 | 31.4 | 11.4 | 332.4 | 299.6 | 60.8% | 68.9% |
| | **100%** | 1546.9 | 28.9 | 10.3 | 311.3 | 313.2 | 64.0% | 71.9% |
| | **RW** | 1404.5 | 31.9 | 11.7 | 334.5 | 297.1 | 60.2% | 68.1% |
| **FIT** | **30%** | 811.4 | 46.7 | 18.2 | 466.4 | 220.2 | 41.8% | 50.5% |
| | **40%** | 972.7 | 40.8 | 15.3 | 424.9 | 254.2 | 49.1% | 58.4% |
| | **50%** | 1102.8 | 36.7 | 13.3 | 391.3 | 278.8 | 54.3% | 63.8% |
| | **60%** | 1229.3 | 33.2 | 11.8 | 361.9 | 295.8 | 58.5% | 68.0% |
| | **70%** | 1321.5 | 30.9 | 10.9 | 339.6 | 308.1 | 61.5% | 70.5% |
| | **100%** | 1585.5 | 25.6 | 8.7 | 286.9 | 333.1 | 68.0% | 76.2% |
| | **RW** | 1004.9 | 39.8 | 14.9 | 419.2 | 258.9 | 50.3% | 59.6% |
| **FOBT** | **30%** | 647.0 | 57.2 | 22.6 | 562.8 | 174.5 | 28.6% | 38.7% |
| | **40%** | 794.6 | 52.1 | 19.7 | 534.4 | 206.4 | 35.0% | 46.5% |
| | **50%** | 923.9 | 47.7 | 17.5 | 507.1 | 232.3 | 40.5% | 52.3% |
| | **60%** | 1040.4 | 44.1 | 15.6 | 485.1 | 256.6 | 44.9% | 57.6% |
| | **70%** | 1148.8 | 40.8 | 14.2 | 457.0 | 273.9 | 49.0% | 61.3% |
| | **100%** | 1426.6 | 33.8 | 11.2 | 392.8 | 306.6 | 57.8% | 69.5% |
| | **RW** | 698.3 | 55.3 | 21.6 | 551.5 | 185.1 | 30.9% | 41.4% |
| **Blood, S1** | **30%** | 820.6 | 60.0 | 24.3 | 576.1 | 151.3 | 25.1% | 34.0% |
| | **40%** | 938.5 | 57.0 | 22.5 | 563.0 | 172.9 | 28.9% | 38.9% |

| | | | | | | | | |
|---|---|---|---|---|---|---|---|---|
| **nAAS=10%<br>AAS=10%** | **50%** | 1030.0 | 54.6 | 21.2 | 548.5 | 188.1 | 31.9% | 42.3% |
| | **60%** | 1099.6 | 53.0 | 20.3 | 541.7 | 199.6 | 33.9% | 44.9% |
| | **70%** | 1157.2 | 51.7 | 19.6 | 535.7 | 208.0 | 35.5% | 46.7% |
| | **100%** | 1285.8 | 48.7 | 17.9 | 519.1 | 227.5 | 39.2% | 51.3% |
| **Blood, S2<br>nAAS=12.5%<br>AAS= 15%** | **30%** | 920.4 | 54.0 | 21.9 | 523.3 | 175.0 | 32.6% | 40.6% |
| | **40%** | 1043.3 | 50.8 | 20.0 | 506.8 | 197.9 | 36.6% | 45.6% |
| | **50%** | 1146.0 | 48.1 | 18.6 | 487.9 | 214.4 | 40.0% | 49.4% |
| | **60%** | 1216.9 | 46.5 | 17.8 | 479.3 | 224.5 | 42.0% | 51.7% |
| | **70%** | 1275.4 | 44.8 | 16.8 | 468.1 | 235.8 | 44.1% | 54.2% |
| | **100%** | 1405.9 | 41.7 | 15.3 | 447.3 | 253.8 | 48.0% | 58.4% |
| **Blood, S3<br>nAAS=15%<br>AAS= 15%** | **30%** | 967.2 | 52.3 | 21.1 | 509.2 | 183.4 | 34.7% | 42.7% |
| | **40%** | 1101.9 | 48.5 | 19.1 | 487.0 | 206.8 | 39.5% | 48.0% |
| | **50%** | 1194.8 | 46.1 | 17.9 | 471.0 | 221.6 | 42.5% | 51.4% |
| | **60%** | 1268.3 | 43.9 | 16.8 | 456.0 | 234.1 | 45.2% | 54.5% |
| | **70%** | 1339.8 | 42.3 | 16.0 | 445.9 | 242.9 | 47.2% | 56.4% |
| | **100%** | 1470.2 | 39.3 | 14.4 | 426.3 | 262.4 | 50.9% | 60.8% |
| **Blood, S4<br>nAAS=15%<br>AAS= 20%** | **30%** | 1003.7 | 49.1 | 19.8 | 479.1 | 196.4 | 38.7% | 46.1% |
| | **40%** | 1134.9 | 45.6 | 17.9 | 460.1 | 219.7 | 43.1% | 51.4% |
| | **50%** | 1239.0 | 43.0 | 16.7 | 439.5 | 234.4 | 46.3% | 54.6% |
| | **60%** | 1316.4 | 40.9 | 15.5 | 424.0 | 247.7 | 49.0% | 57.8% |
| | **70%** | 1369.6 | 39.5 | 14.9 | 414.3 | 256.3 | 50.7% | 59.4% |
| | **100%** | 1505.7 | 36.4 | 13.4 | 391.7 | 272.9 | 54.5% | 63.7% |
| **Blood, S5<br>nAAS=20%<br>AAS= 20%** | **30%** | 1089.2 | 45.5 | 18.2 | 451.3 | 212.5 | 43.2% | 50.5% |
| | **40%** | 1231.3 | 41.8 | 16.4 | 425.7 | 234.1 | 47.8% | 55.5% |
| | **50%** | 1334.8 | 39.1 | 15.0 | 407.3 | 250.4 | 51.2% | 59.2% |
| | **60%** | 1409.7 | 37.3 | 14.2 | 392.4 | 261.4 | 53.5% | 61.5% |
| | **70%** | 1472.6 | 35.7 | 13.4 | 380.2 | 270.2 | 55.4% | 63.5% |
| | **100%** | 1601.9 | 32.7 | 11.9 | 358.2 | 287.2 | 59.2% | 67.6% |

AAS, advanced adenoma (≥ 10 mm) sensitivity; COL, colonoscopy; CRC, colorectal cancer; FIT, fecal immunochemical test; FOBT, fecal occult blood test; LY, life-years; LYG, life-years gained; mt-sDNA, multi-target stool DNA; nAAS, non-advanced adenoma (<10 mm) sensitivity; RW, real-world; S, scenario.

**Table S2.** Estimated outcomes of sensitivity analyses with blood-based test under various adherence and expanded blood-based test adenoma sensitivity scenarios.

| Blood-test sensitivity | Adherence | Total COLs | CRC cases | CRC deaths | LYG | CRC Incidence reduction | CRC Mortality reduction |
|---|---|---|---|---|---|---|---|
| **nAAS=20% AAS= 30%** | 30% | 1147.4 | 41.4 | 16.5 | 231.3 | 48.3% | 55.1% |
| | 40% | 1279.0 | 37.7 | 14.7 | 252.6 | 53.0% | 59.9% |
| | 50% | 1382.9 | 35.0 | 13.5 | 268.5 | 56.4% | 63.3% |
| | 60% | 1462.4 | 33.0 | 12.6 | 278.8 | 58.8% | 65.8% |
| | 70% | 1521.6 | 31.6 | 11.9 | 287.4 | 60.6% | 67.7% |
| | 100% | 1647.9 | 29.0 | 10.9 | 300.7 | 63.8% | 70.5% |
| **nAAS=30% AA= 30%** | 30% | 1288.7 | 36.1 | 14.4 | 253.1 | 54.9% | 60.8% |
| | 40% | 1426.9 | 32.3 | 12.6 | 272.6 | 59.7% | 65.7% |
| | 50% | 1526.3 | 29.9 | 11.5 | 287.9 | 62.7% | 68.8% |
| | 60% | 1602.3 | 28.0 | 10.7 | 298.4 | 65.0% | 71.0% |
| | 70% | 1665.2 | 26.9 | 10.1 | 305.6 | 66.4% | 72.6% |
| | 100% | 1788.0 | 24.6 | 9.1 | 317.9 | 69.3% | 75.2% |
| **nAAS=25% AAS= 40%** | 30% | 1263.3 | 35.3 | 14.2 | 256.7 | 55.9% | 61.5% |
| | 40% | 1399.2 | 31.7 | 12.4 | 278.6 | 60.4% | 66.3% |
| | 50% | 1499.4 | 29.2 | 11.2 | 293.3 | 63.5% | 69.5% |
| | 60% | 1568.1 | 27.8 | 10.7 | 300.3 | 65.3% | 71.0% |
| | 70% | 1629.4 | 26.7 | 10.1 | 307.5 | 66.7% | 72.6% |
| | 100% | 1747.3 | 24.5 | 9.2 | 319.8 | 69.4% | 75.1% |
| **nAAS=40% AAS= 40%** | 30% | 1430.8 | 29.5 | 11.8 | 281.9 | 63.2% | 68.1% |
| | 40% | 1563.1 | 26.2 | 10.1 | 301.7 | 67.3% | 72.4% |
| | 50% | 1662.6 | 23.9 | 9.2 | 313.7 | 70.1% | 74.9% |
| | 60% | 1731.1 | 22.6 | 8.6 | 320.3 | 71.8% | 76.5% |
| | 70% | 1790.6 | 21.8 | 8.2 | 325.9 | 72.8% | 77.6% |
| | 100% | 1905.7 | 20.3 | 7.5 | 336.7 | 74.7% | 79.5% |

AAS, advanced adenoma (≥ 10 mm) sensitivity; COL, colonoscopy; CRC, colorectal cancer; LYG, life-years gained; nAAS, non-advanced adenoma (<10 mm) sensitivity.

**Figure S1.** Colorectal cancer incidence reduction (IR) per 1,000 patients screened over lifetime horizon based on different adherence rates blood-based test adenoma sensitivity at A) 10% and B) 20%. Numbers represent ranges of incremental IR versus the blood-based test. A positive value (green) indicates the stool-based test has less IR than the blood-based test, and a negative value (red/orange) indicates the stool-based test has more IR than the blood-based test. Adherence rates ranged from 100% or 30-70% for both stool- and blood-tests. FIT, fecal immunochemical test; FOBT, fecal occult blood test; mt-sDNA, multi-target stool DNA.

A)

| | | mt-sDNA | | | | | | |
|---|---|---|---|---|---|---|---|---|
| | Screening adherence rate | 30% | 40% | 50% | 60% | 70% | 100% | Blood IR |
| Blood Test | 30% | -25% | -29% | -32% | -34% | -36% | -39% | 25% |
| | 40% | -21% | -25% | -28% | -30% | -32% | -35% | 29% |
| | 50% | -18% | -22% | -25% | -27% | -29% | -32% | 32% |
| | 60% | -16% | -20% | -23% | -25% | -27% | -30% | 34% |
| | 70% | -14% | -19% | -22% | -24% | -25% | -29% | 35% |
| | 100% | -11% | -15% | -18% | -20% | -22% | -25% | 39% |
| | mt-sDNA IR | 50% | 54% | 57% | 59% | 61% | 64% | |

| | | FIT | | | | | | |
|---|---|---|---|---|---|---|---|---|
| | Screening adherence rate | 30% | 40% | 50% | 60% | 70% | 100% | Blood IR |
| Blood Test | 30% | -17% | -24% | -29% | -33% | -36% | -43% | 25% |
| | 40% | -13% | -20% | -25% | -30% | -33% | -39% | 29% |
| | 50% | -10% | -17% | -22% | -27% | -30% | -36% | 32% |
| | 60% | -8% | -15% | -20% | -25% | -28% | -34% | 34% |
| | 70% | -6% | -14% | -19% | -23% | -26% | -33% | 35% |
| | 100% | -3% | -10% | -15% | -19% | -22% | -29% | 39% |
| | FIT IR | 42% | 49% | 54% | 59% | 61% | 68% | |

| | | FOBT | | | | | | |
|---|---|---|---|---|---|---|---|---|
| | Screening adherence rate | 30% | 40% | 50% | 60% | 70% | 100% | Blood IR |
| Blood Test | 30% | -4% | -10% | -15% | -20% | -24% | -33% | 25% |
| | 40% | 0% | -6% | -12% | -16% | -20% | -29% | 29% |
| | 50% | 3% | -3% | -9% | -13% | -17% | -26% | 32% |
| | 60% | 5% | -1% | -7% | -11% | -15% | -24% | 34% |
| | 70% | 7% | 0% | -5% | -9% | -14% | -22% | 35% |
| | 100% | 11% | 4% | -1% | -6% | -10% | -19% | 39% |
| | FOBT IR | 29% | 35% | 41% | 45% | 49% | 58% | |

B)

| | | mt-sDNA | | | | | | |
|---|---|---|---|---|---|---|---|---|
| | Screening adherence rate | 30% | 40% | 50% | 60% | 70% | 100% | Blood IR |
| Blood Test | 30% | -7% | -11% | -14% | -16% | -18% | -21% | 43% |
| | 40% | -2% | -6% | -9% | -11% | -13% | -16% | 48% |
| | 50% | 1% | -3% | -6% | -8% | -10% | -13% | 51% |
| | 60% | 4% | -1% | -4% | -6% | -7% | -10% | 53% |
| | 70% | 5% | 1% | -2% | -4% | -5% | -9% | 55% |
| | 100% | 9% | 5% | 2% | 0% | -2% | -5% | 59% |
| | mt-sDNA IR | 50% | 54% | 57% | 59% | 61% | 64% | |

| | | FIT | | | | | | |
|---|---|---|---|---|---|---|---|---|
| | Screening adherence rate | 30% | 40% | 50% | 60% | 70% | 100% | Blood IR |
| Blood Test | 30% | 1% | -6% | -11% | -15% | -18% | -25% | 43% |
| | 40% | 6% | -1% | -6% | -11% | -14% | -20% | 48% |
| | 50% | 9% | 2% | -3% | -7% | -10% | -17% | 51% |
| | 60% | 12% | 4% | -1% | -5% | -8% | -15% | 53% |
| | 70% | 14% | 6% | 1% | -3% | -6% | -13% | 55% |
| | 100% | 17% | 10% | 5% | 1% | -2% | -9% | 59% |
| | FIT IR | 42% | 49% | 54% | 59% | 61% | 68% | |

| | | FOBT | | | | | | |
|---|---|---|---|---|---|---|---|---|
| | Screening adherence rate | 30% | 40% | 50% | 60% | 70% | 100% | Blood IR |
| Blood Test | 30% | 15% | 8% | 3% | -2% | -6% | -15% | 43% |
| | 40% | 19% | 13% | 7% | 3% | -1% | -10% | 48% |
| | 50% | 23% | 16% | 11% | 6% | 2% | -7% | 51% |
| | 60% | 25% | 18% | 13% | 9% | 4% | -4% | 53% |
| | 70% | 27% | 20% | 15% | 10% | 6% | -2% | 55% |
| | 100% | 31% | 24% | 19% | 14% | 10% | 1% | 59% |
| | FOBT IR | 29% | 35% | 41% | 45% | 49% | 58% | |

**Figure S2.** Colorectal cancer mortality reduction (MR) per 1,000 patients screened over lifetime horizon based on different adherence rates for blood-based test adenoma sensitivity at A) 10% and B) 20%. Numbers represent ranges of incremental MR versus the blood-based test. A positive value (green) indicates the stool-based test has less MR than the blood-based test, and a negative value (red/orange) indicates the stool-based test has more MR than the blood-based test. Adherence rates ranged from 100% or 30-70% for both stool- and blood-tests. FIT, fecal immunochemical test; FOBT, fecal occult blood test; mt-sDNA, multi-target stool DNA.

## A)

| | | mt-sDNA | | | | | | |
|---|---|---|---|---|---|---|---|---|
| | Screening adherence rate | 30% | 40% | 50% | 60% | 70% | 100% | Blood MR |
| Blood Test | 30% | -24% | -28% | -31% | -33% | -35% | -38% | 34% |
| | 40% | -19% | -23% | -26% | -29% | -30% | -33% | 39% |
| | 50% | -16% | -20% | -23% | -25% | -27% | -30% | 42% |
| | 60% | -13% | -17% | -20% | -22% | -24% | -27% | 45% |
| | 70% | -11% | -16% | -19% | -21% | -22% | -25% | 47% |
| | 100% | -7% | -11% | -14% | -16% | -18% | -21% | 51% |
| | mt-sDNA MR | 58% | 62% | 65% | 67% | 69% | 72% | |

| | | FIT | | | | | | |
|---|---|---|---|---|---|---|---|---|
| | Screening adherence rate | 30% | 40% | 50% | 60% | 70% | 100% | Blood MR |
| Blood Test | 30% | -17% | -24% | -30% | -34% | -36% | -42% | 34% |
| | 40% | -12% | -19% | -25% | -29% | -32% | -37% | 39% |
| | 50% | -8% | -16% | -22% | -26% | -28% | -34% | 42% |
| | 60% | -6% | -13% | -19% | -23% | -26% | -31% | 45% |
| | 70% | -4% | -12% | -17% | -21% | -24% | -30% | 47% |
| | 100% | 1% | -7% | -13% | -17% | -19% | -25% | 51% |
| | FIT MR | 50% | 58% | 64% | 68% | 70% | 76% | |

| | | FOBT | | | | | | |
|---|---|---|---|---|---|---|---|---|
| | Screening adherence rate | 30% | 40% | 50% | 60% | 70% | 100% | Blood MR |
| Blood Test | 30% | -5% | -12% | -18% | -24% | -27% | -35% | 34% |
| | 40% | 0% | -8% | -13% | -19% | -22% | -31% | 39% |
| | 50% | 4% | -4% | -10% | -15% | -19% | -27% | 42% |
| | 60% | 6% | -2% | -7% | -13% | -16% | -25% | 45% |
| | 70% | 8% | 0% | -6% | -11% | -15% | -23% | 47% |
| | 100% | 13% | 5% | -1% | -6% | -10% | -18% | 51% |
| | FOBT MR | 39% | 46% | 52% | 58% | 61% | 69% | |

## B)

| | | mt-sDNA | | | | | | |
|---|---|---|---|---|---|---|---|---|
| | Screening adherence rate | 30% | 40% | 50% | 60% | 70% | 100% | Blood MR |
| Blood Test | 30% | -7% | -12% | -15% | -17% | -18% | -21% | 50% |
| | 40% | -2% | -7% | -10% | -12% | -13% | -16% | 55% |
| | 50% | 1% | -3% | -6% | -8% | -10% | -13% | 59% |
| | 60% | 4% | -1% | -4% | -6% | -7% | -10% | 62% |
| | 70% | 6% | 1% | -2% | -4% | -5% | -8% | 63% |
| | 100% | 10% | 5% | 2% | 0% | -1% | -4% | 68% |
| | mt-sDNA MR | 58% | 62% | 65% | 67% | 69% | 72% | |

| | | FIT | | | | | | |
|---|---|---|---|---|---|---|---|---|
| | Screening adherence rate | 30% | 40% | 50% | 60% | 70% | 100% | Blood MR |
| Blood Test | 30% | 0% | -8% | -13% | -17% | -20% | -26% | 50% |
| | 40% | 5% | -3% | -8% | -12% | -15% | -21% | 55% |
| | 50% | 9% | 1% | -5% | -9% | -11% | -17% | 59% |
| | 60% | 11% | 3% | -2% | -6% | -9% | -15% | 62% |
| | 70% | 13% | 5% | 0% | -4% | -7% | -13% | 63% |
| | 100% | 17% | 9% | 4% | 0% | -3% | -9% | 68% |
| | FIT MR | 50% | 58% | 64% | 68% | 70% | 76% | |

| | | FOBT | | | | | | |
|---|---|---|---|---|---|---|---|---|
| | Screening adherence rate | 30% | 40% | 50% | 60% | 70% | 100% | Blood MR |
| Blood Test | 30% | 12% | 4% | -2% | -7% | -11% | -19% | 50% |
| | 40% | 17% | 9% | 3% | -2% | -6% | -14% | 55% |
| | 50% | 20% | 13% | 7% | 2% | -2% | -10% | 59% |
| | 60% | 23% | 15% | 9% | 4% | 0% | -8% | 62% |
| | 70% | 25% | 17% | 11% | 6% | 2% | -6% | 63% |
| | 100% | 29% | 21% | 15% | 10% | 6% | -2% | 68% |
| | FOBT MR | 39% | 46% | 52% | 58% | 61% | 69% | |
